# Concerning Pregnancy Weight Trends by Race and Ethnicity in the United States, 2016–2023

**DOI:** 10.1101/2025.10.15.25338088

**Authors:** Amos Grünebaum, Joachim Dudenhausen, Frank A. Chervenak

## Abstract

**Background:** Maternal prepregnancy body mass index (BMI) is a critical determinant of pregnancy outcomes, influencing risks for gestational diabetes, hypertensive disorders, cesarean delivery, stillbirth, and long-term maternal and child health. Although the overall rise in obesity among reproductive-aged women in the United States is well recognized, recent national data describing detailed trends by race and ethnicity are limited.

**Objective:** To examine temporal trends in prepregnancy BMI among U.S. mothers from 2016 to 2023, with a focus on changes in normal weight and obesity prevalence across racial and ethnic groups.

**Methods:** We performed a retrospective analysis of nearly 30 million live births recorded in the CDC Natality database. Maternal BMI was categorized as underweight, normal weight, overweight, and obesity classes I–III. Race and ethnicity were defined using CDC conventions: non-Hispanic (NH) White, NH Black, NH Asian, NH American Indian or Alaska Native, NH Native Hawaiian or Pacific Islander, NH more than one race, and Hispanic (all races). Annual prevalence for each BMI category was calculated, and linear regression was used to estimate slopes of annual change with 95% confidence intervals.

**Results:** Nationally, normal weight prevalence declined from 42% in 2016 to 37% in 2023, while combined obesity increased from 24% to 29%. Declines in normal weight and corresponding increases in obesity were observed across all racial and ethnic groups. Asian mothers had the highest prevalence of normal weight (60% in 2016, 53% in 2023) and the lowest obesity prevalence (<15%), yet still showed significant annual increases. By 2023, obesity prevalence exceeded 30% among Black and American Indian/Alaska Native mothers and reached 25–30% among Hispanic and multiracial mothers. Hispanic and Black mothers experienced the steepest annual increases in obesity (+0.98% and +0.93% per year, respectively). Projections indicate that normal weight prevalence could fall below 30% within the next decade, while obesity prevalence may approach 40% by the mid-2030s.

**Conclusions:** Between 2016 and 2023, U.S. mothers experienced a population-wide redistribution from normal weight to obesity, affecting all racial and ethnic groups. These trends pose substantial risks for adverse pregnancy outcomes and threaten to exacerbate maternal health inequities. Equity-focused preconception counseling, nutrition education, and policy-level interventions are urgently needed.

## Introduction

Maternal prepregnancy body mass index (BMI) is a critical determinant of pregnancy outcomes, influencing risks for gestational diabetes, hypertensive disorders, cesarean delivery, stillbirth, and long-term maternal and child health. Over the past two decades, the prevalence of overweight and obesity among women of reproductive age in the United States has risen sharply, raising concern for both clinical practice and public health. National surveillance is essential for understanding how these trends evolve and whether disparities by race and ethnicity are widening.

The Centers for Disease Control and Prevention (CDC) Natality database provides a unique opportunity to assess prepregnancy BMI across all live births in the United States. Although the overall rise in obesity is well recognized, there has been limited documentation of year-to-year changes across detailed BMI categories and their distribution by race and ethnicity in recent years. Understanding these dynamics is critical not only for epidemiologic surveillance but also for guiding healthcare systems and policy. Rising maternal obesity has direct implications for the allocation of obstetric resources, the design of prenatal counseling programs, and the development of preventive strategies aimed at reducing adverse outcomes.

## Objective

The objective of this study was to analyze trends in prepregnancy BMI among U.S. mothers from 2016 to 2023 using the CDC Natality database. The study aimed to quantify the annual prevalence of underweight, normal weight, overweight, and obesity classes I–III by race and ethnicity, to calculate slopes of annual change with 95% confidence intervals to assess the statistical significance of temporal trends, and to identify disparities in both baseline prevalence and rates of change across racial and ethnic groups. By linking these findings to policy and healthcare practice, the goal was to provide actionable evidence to inform targeted interventions, reduce inequities, and anticipate future needs in obstetric care.

## Methods

We conducted a retrospective analysis using the publicly available **CDC Natality database** for the years **2016–2023**. The dataset includes all live births in the United States as reported on birth certificates. Maternal pre-pregnancy body mass index (BMI) was categorized by the CDC into the following groups:

- **Underweight:** BMI <18.5
- **Normal weight:** BMI 18.5–24.9
- **Overweight:** BMI 25.0–29.9
- **Obesity I:** BMI 30.0–34.9
- **Obesity II:** BMI 35.0–39.9
- **Obesity III:** BMI ≥40.0

Records with missing BMI data were retained in the **Unknown** category.

Race and ethnicity were derived from birth certificate reporting. Consistent with CDC conventions, Hispanic women were classified as Hispanic regardless of race, while all other race categories (White, Black, Asian, American Indian or Alaska Native, and women reporting more than one race) were limited to non-Hispanic (NH) women or those with unknown Hispanic ethnicity.

For each weight group, we calculated the annual prevalence of live births by maternal race/ethnicity from 2016 through 2023. Linear regression was used to estimate slopes of annual change in prevalence (percentage points per year) along with 95% confidence intervals (CI). Separate models were fit for each race/ethnic group and weight category, with year as the independent variable.

## Results

### Overall Trends

From 2016 to 2023, normal weight prevalence declined from 42% to 37% nationally, while combined obesity (classes I-III) increased from 24% to 29%. These trends occurred across all racial and ethnic groups, demonstrating a population-wide redistribution from normal weight to obesity categories.

### Racial and Ethnic Disparities

#### Underweight

All groups showed declining prevalence. Asian mothers had the highest levels, falling from 8% to 5%, while most other groups remained at 2-3%.

#### Normal Weight

Asian mothers maintained the highest prevalence but declined from 60% to 53%. Non-Hispanic White mothers decreased from 47% to 41%. Hispanic, Black, and multiracial mothers fell into the 20-30% range, while American Indian/Alaska Native and Pacific Islander mothers reached the lowest levels at 20-27% by 2023.

#### Overweight

Prevalence remained relatively stable across the study period. Hispanic mothers consistently showed the highest rates (∼30%), followed by other groups at 25-27%. Asian mothers increased from 20% to 24%.

#### Combined Obesity (Classes I-III)

By 2023, Black and American Indian/Alaska Native mothers exceeded 30% obesity prevalence. Hispanic and multiracial mothers reached 25-30%, while White mothers were at 20-25%. Asian mothers remained lowest at <15% but showed significant increases.

### Annual Rates of Change

Linear regression analysis revealed statistically significant trends across all BMI categories. For combined obesity, Hispanic mothers showed the steepest annual increase at +0.98% per year (95% CI 0.80 to 1.15), followed by Black mothers at +0.93% (95% CI 0.78 to 1.08). American Indian/Alaska Native mothers increased by +0.80% annually (95% CI 0.66 to 0.94), multiracial mothers by +0.86% (95% CI 0.73 to 1.00), and Asian mothers by +0.79% (95% CI 0.74 to 0.85).

These obesity increases were mirrored by significant declines in normal weight prevalence. Asian and Hispanic mothers showed the steepest decreases at -1.04% and -1.06% per year, respectively, while American Indian/Alaska Native and Black mothers declined at -0.69% and - 0.70% annually. All confidence intervals excluded zero, confirming statistical significance.

Individual obesity classes showed consistent upward trends. Obesity Class I increased most rapidly among American Indian/Alaska Native mothers (+0.227% annually), while Obesity Classes II and III showed the steepest rises among Hispanic mothers (+0.291% and +0.195% annually, respectively).

## Discussion

This national analysis of nearly 30 million births demonstrates a concerning redistribution of maternal prepregnancy BMI in the United States from 2016 to 2023. Normal weight prevalence declined steadily, while obesity increased across all racial and ethnic groups. These findings confirm that the obesity epidemic is firmly established in women of reproductive age, with systemic rather than group-specific drivers.

Disparities are both persistent and widening. By 2023, Black and American Indian/Alaska Native mothers exceeded 30% obesity prevalence, while Hispanic mothers had the steepest annual increases. Asian mothers retained the lowest prevalence but experienced significant year-to-year gains. These patterns parallel longstanding inequities in maternal morbidity and mortality, raising concern that maternal obesity will increasingly contribute to poor outcomes in already vulnerable populations.

The decline in normal weight prevalence has profound clinical implications. Maternal obesity is strongly associated with gestational diabetes, preeclampsia, cesarean delivery, and stillbirth, as well as long-term cardiometabolic risks for both mother and child. Importantly, if the observed linear trends continue, normal weight prevalence could fall below 30% within the next decade, while obesity could approach 40% by the mid-2030s. Such projections underscore that the majority of pregnant women may soon begin pregnancy at elevated risk, further straining obstetric systems and worsening outcome disparities.

At the policy level, universal interventions such as nutrition education, improved food access, and public health messaging should be combined with targeted approaches for communities experiencing the steepest increases. Training clinicians in evidence-based weight counseling and culturally tailored strategies will be critical to translating surveillance data into improved care.

## Conclusion

In summary, maternal obesity in the United States is rising across all racial and ethnic groups, accompanied by a steady decline in normal weight. These trends reflect systemic drivers and portend worsening disparities in maternal and neonatal health if left unaddressed. While the strengths of this study provide confidence in the robustness of the findings, its limitations underscore the need for complementary research that incorporates socioeconomic and structural determinants of health. Addressing maternal obesity is both a clinical and public health priority, requiring coordinated interventions that integrate prevention, equity, and accountability

## Data Availability

All data produced in the present study are available upon reasonable request to the authors

https://wonder.cdc.gov/

---

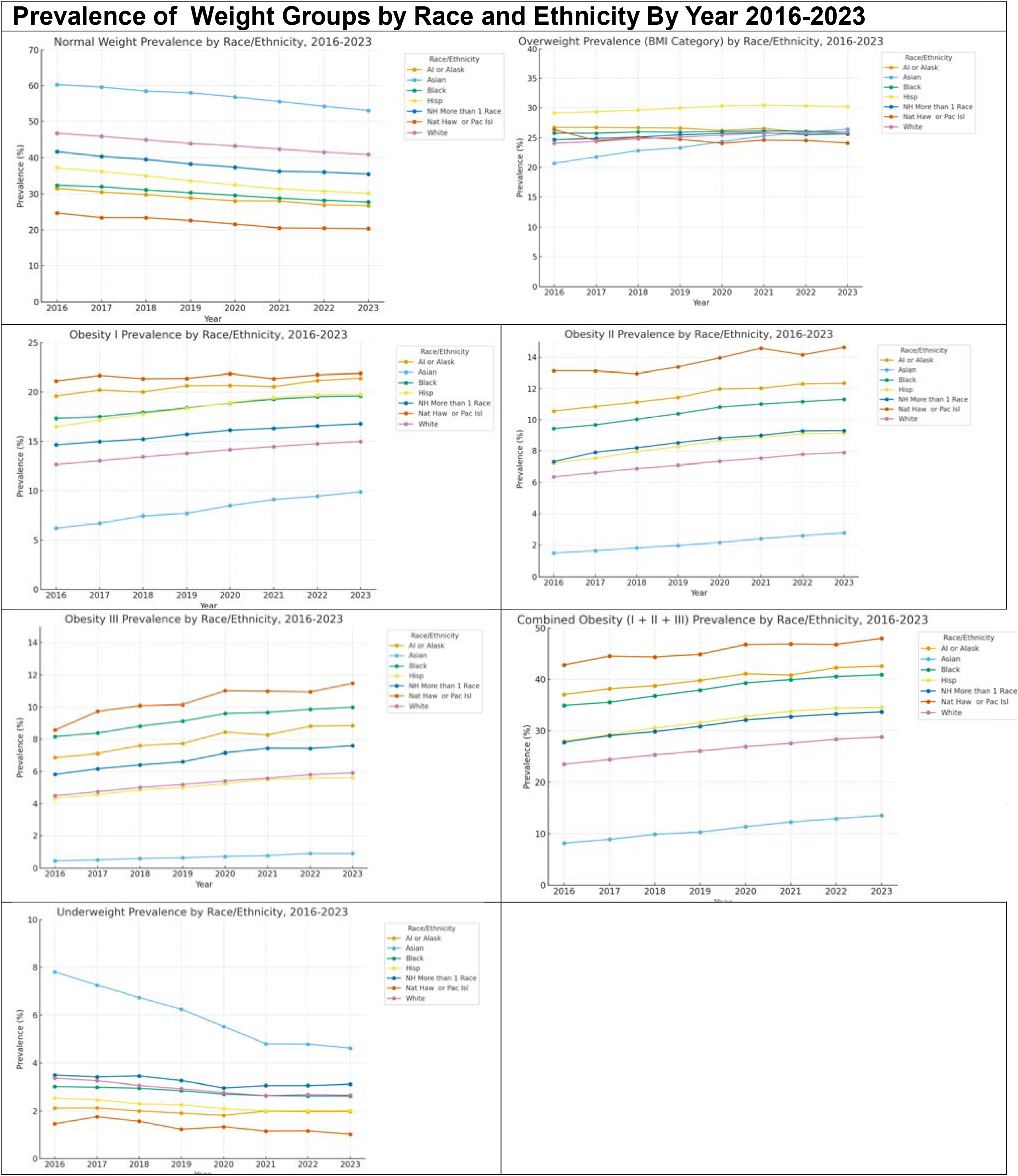

**Table:**
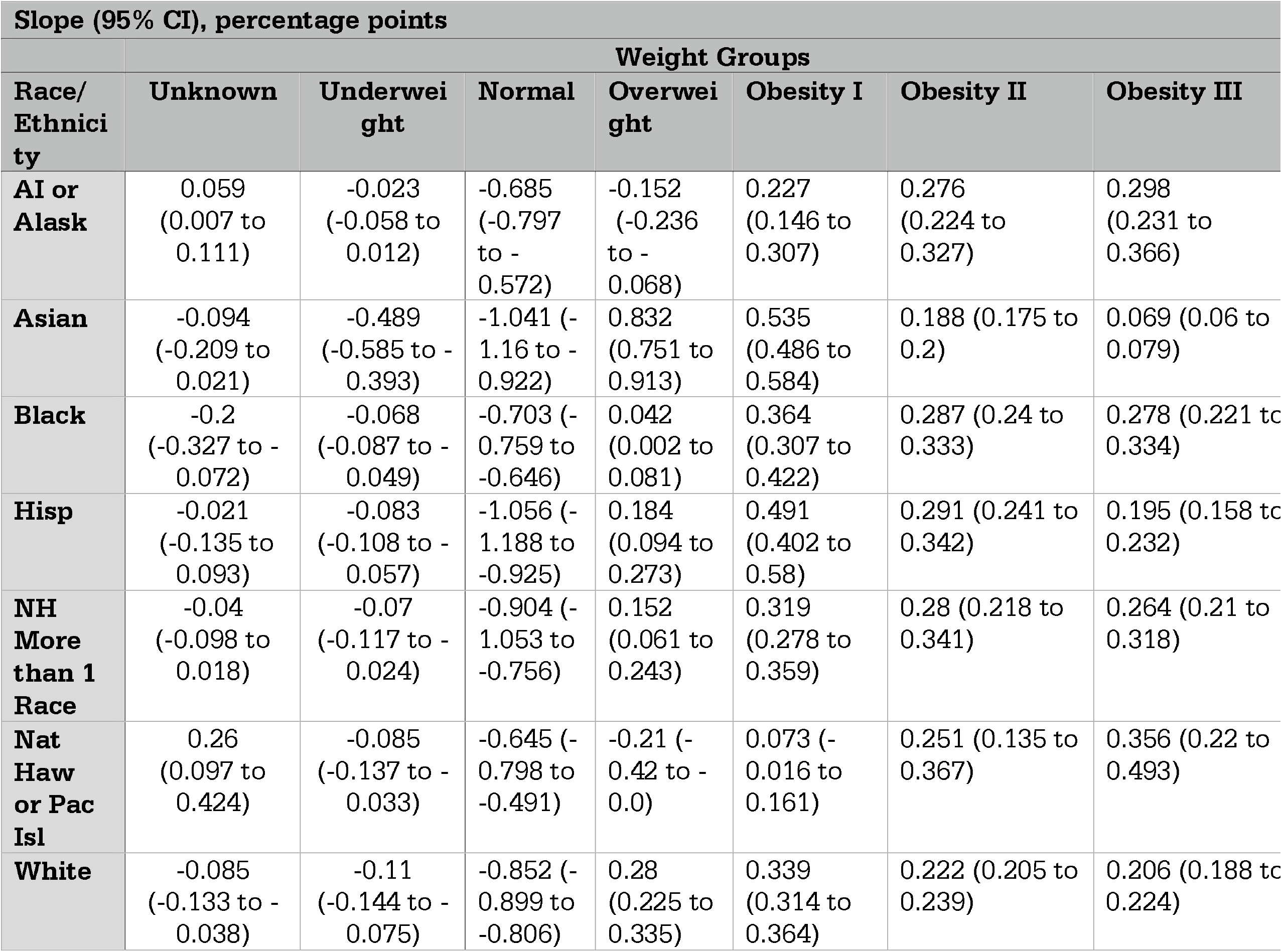
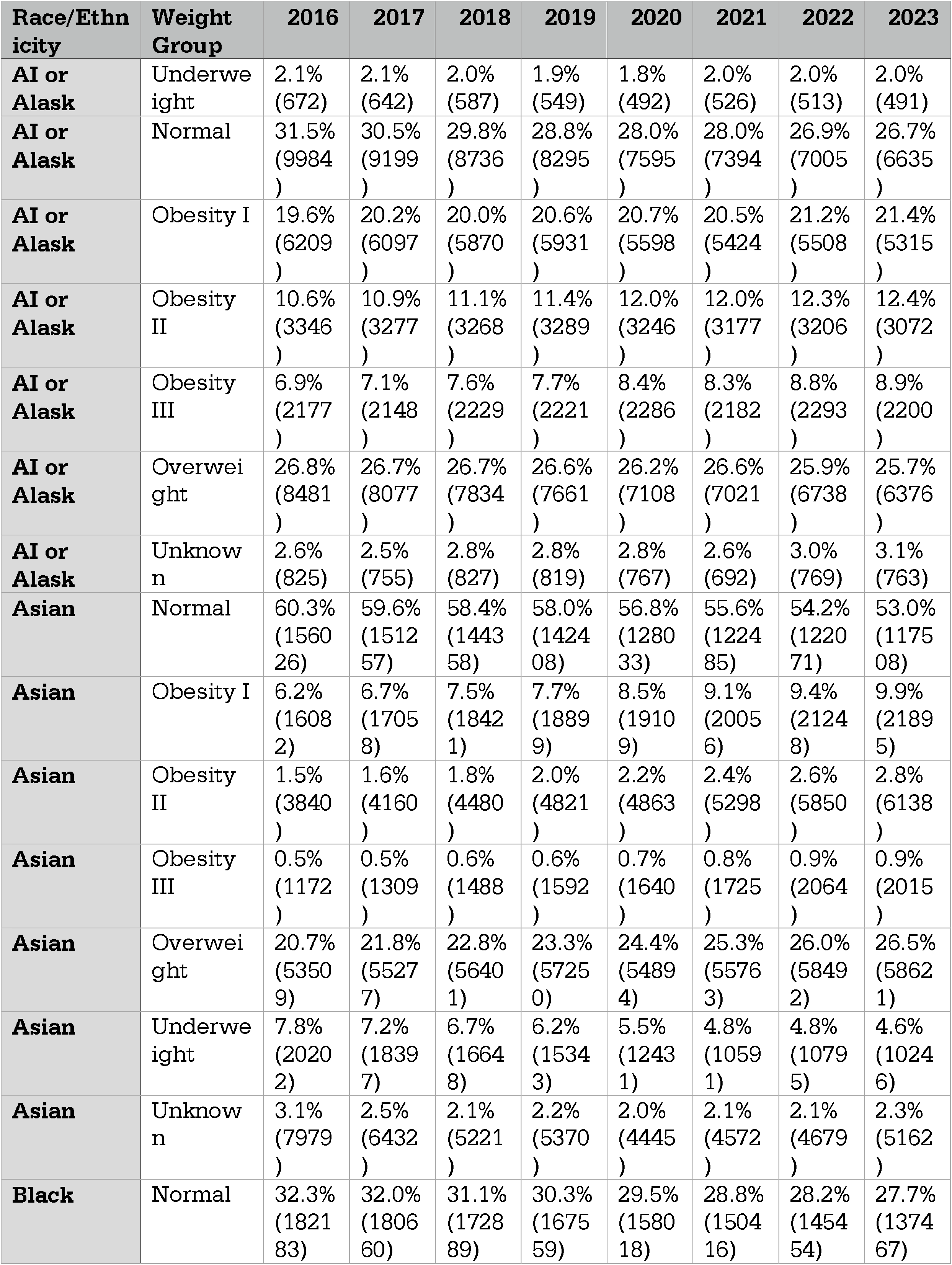
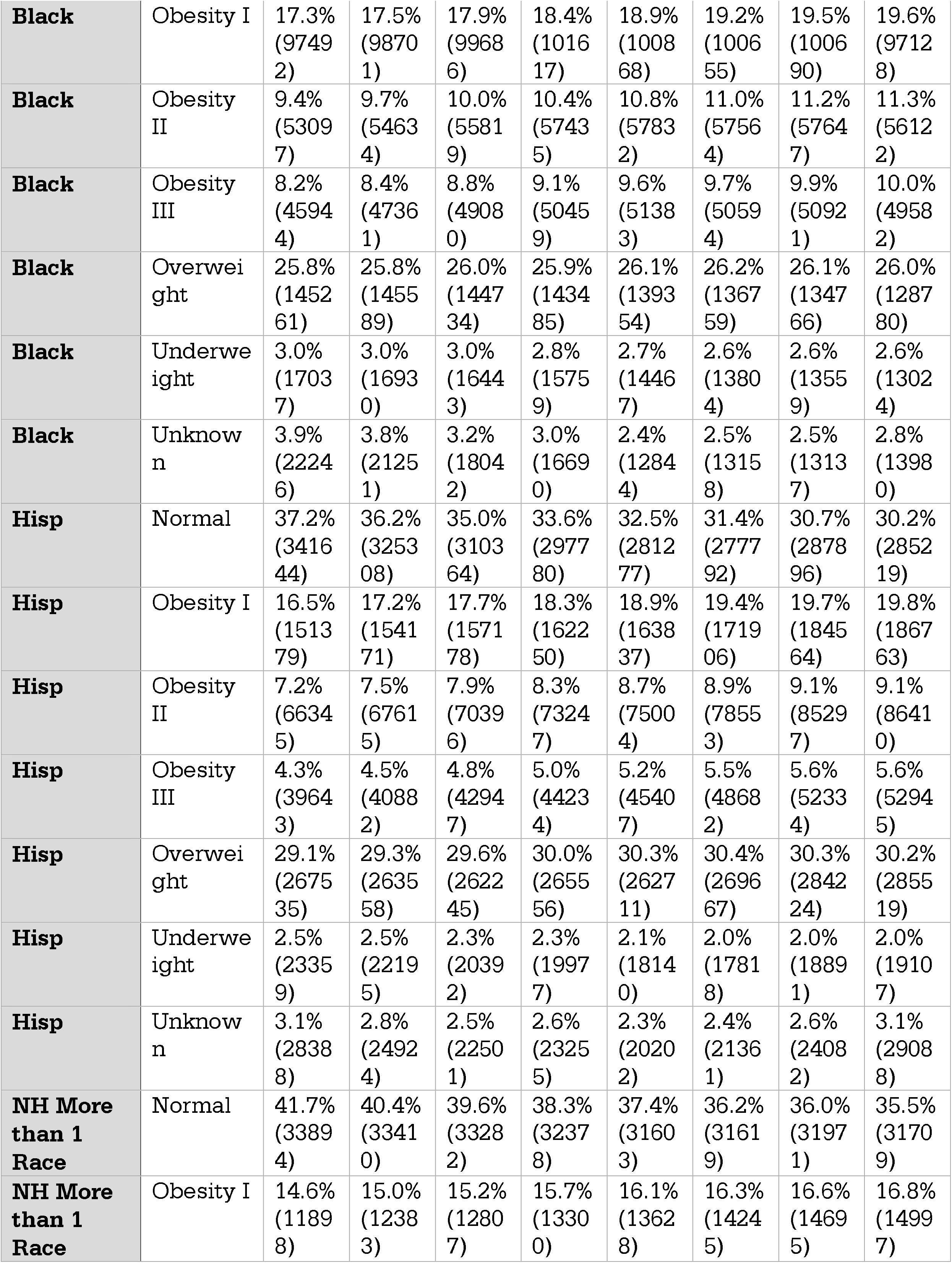
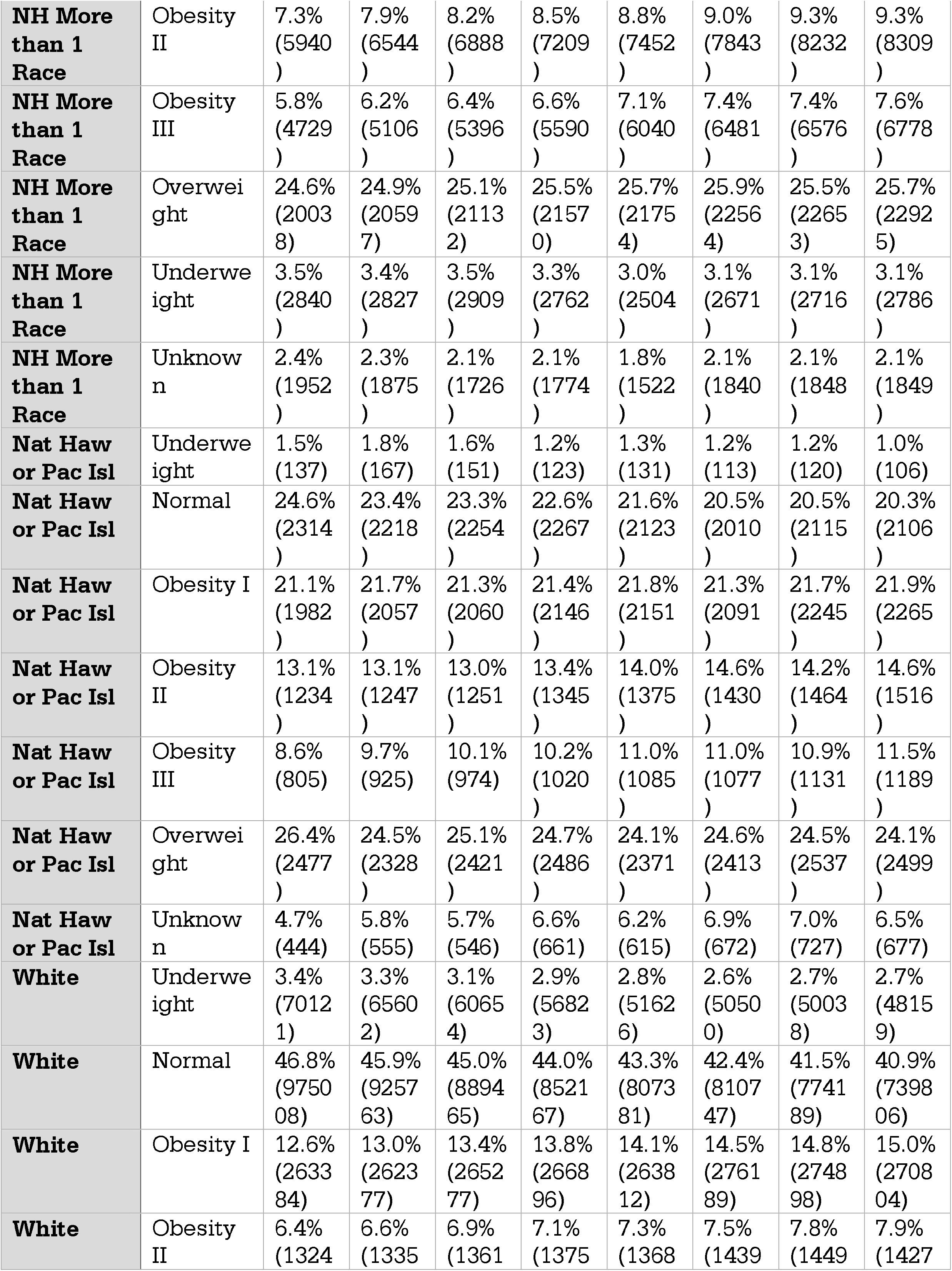
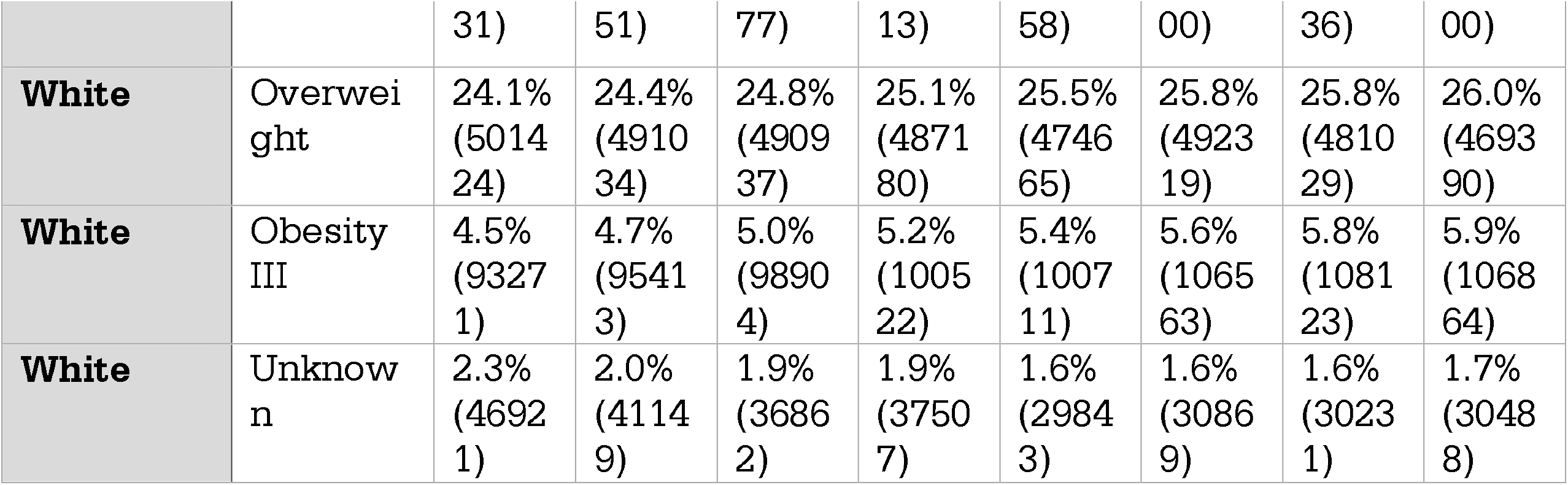
Linear Increase (Slope) of prepregnancy weights per Year by Race and Ethnicity and Prevalence 2016-2023.

